# Emergency Medical Service Response Times and Fatal Fall Injuries Among US Older Adults: Analysis of the 2015 – 2020 National Trauma Data Bank

**DOI:** 10.1101/2023.06.18.23291570

**Authors:** Oluwaseun Adeyemi, Charles DiMaggio, Corita Grudzen, Cuthel Allison, Kaitlyn Van Allen, Joshua Chodosh

## Abstract

**Background:** Falls are the leading injury-related cause of death among older adults but rapid emergency care may reduce fatal complications. To estimate the strength of the association between EMS response times and fatal fall injuries among older adults and measure how this association differs by sex.

**Methods:** For this retrospective cohort study, we pooled 2015 – 2020 data from the National Trauma Data Bank on patients 65 years and older with fall injuries transferred to U.S. trauma centers (N=705,491). The main outcome was fatal fall injuries. The main predictor was EMS response time, measured as continuous and four-level categorical variables (multiples of the standard nine-minute benchmark). Age, sex, race/ethnicity, and diagnoses of COPD, diabetes, and hypertension were covariables. We performed a mixed-effect multivariable logistic regression, using the trauma center designation level as a random effect variable, and EMS response time and mortality as fixed effect variables. We report the unadjusted and adjusted odds ratio (AOR) plus 95% confidence intervals (CI). We also created an interaction model comprising of response time and sex and reported the predicted probabilities (plus 95% CI) of fatal fall injury by sex and response time categories.

**Results:** The case fatality rate of fatal fall injuries among adults 65 years and older was 4.4%. The median (Q1, Q3) EMS response time was 8 minutes (5.0, 13.0), with 60% of patients experiencing the nine-minute benchmark. In the adjusted model, a minute delay in EMS response time was associated with a 1% increased odds of fatal fall injury (AOR: 1.01; 95% CI: 1.01 – 1.01). Older adults who experienced a response time between 18 and 27 minutes, and more than 27 minutes had 1.33 (95% CI: 1.28 - 1.39), and 1.41 (95% CI: 1.35 - 1.47) times the odds of fatal fall injuries. The predicted probabilities of male and female fatal fall injuries were 5.1% (95% CI: 3.51 – 6.75) and 2.4% (95% CI: 1.64 – 3.23), respectively.

**Conclusion:** Delayed EMS response time, especially when above twice the standard benchmark, is associated with increased odds of fatal fall injuries among older adults.

**Key Points:** 

**Question:** What is the association between EMS response time and fatal fall injuries among US older adults?

**Findings:** In this retrospective cohort analysis, delay in EMS response time was associated with fatal fall injury in a dose-response pattern among male and female older adults.

**Meaning:** Strengthening the EMS infrastructure may improve outcomes from fatal fall injuries among older adults.

## Introduction

In the United States (US), falls are the leading cause of injury-related deaths among adults 65 years and older.^1^ Every year, one in four older adults falls, and 20 percent of such falls result in serious injuries such as hip fractures and head injuries.^2, 3^ Fall injuries account for over 32,000 deaths yearly, three million emergency department visits, and 800,000 hospitalizations.^2^ As of 2016, the age-adjusted fatal fall injury rate was 16 per 100,000 among adults 65 to 74 years and 258 per 100,000 among adults 85 years and older.^4^ While falls occur more commonly among women,^5^ the age-adjusted fatal fall injuries are 30 percent higher among men compared to women.^4^

The Emergency Medical Service (EMS), a specialized unit of highly skilled healthcare providers trained in the art of delivering pre-hospital care to acutely and critically ill patients,^6^ is often the immediate healthcare access point among older adults with fall injuries. Earlier studies have reported that falls accounted for 17% of EMS calls involving older adults with approximately 80 to 85 percent requiring transport.^7, 8^ Compared to adults 65 years and older, those aged 85 years and older were more likely to be transported to levels I and II trauma centers.^7, 8^ Also, women and those who sustain fall injuries in residential institutions or healthcare facilities were more likely to be transported by the EMS.^7, 8^ While patient transportation is a critical service delivered by EMS, EMS workers also provide life-saving interventions, which makes such pre-hospital care a pivotal part of critical and acute care delivery.

EMS response can be conceptualized as occurring in four non-overlapping phases: from injury occurrence to EMS notification, EMS notification to chute initiation, chute initiation to arrival at the injury scene, and injury scene to hospital arrival.^9, 10^ Earlier studies have reported that the duration from chute initiation to arrival at the injury scene is the most predictive measure of decreased injury fatality.^11, 12^ Also, a minute increase in the time to arrive at the site where care is needed has been associated with increased odds of death from myocardial infarction,^13^ drowning,^14^ deaths at motor vehicle crash scene,^9^ and county-level crash mortality rate ratios.^11, 12^ No study has explored the relationship between any of the phases of EMS response times and fatal fall injuries.

Since falls account for over 70 percent of injuries among older adults^15^ and over 10,000 US adults turn 65 years daily,^16^ understanding the relationship between EMS response and fatal fall injuries is essential. The knowledge of the relationship between fall injuries and EMS response in older adults may provide information that will strengthen the EMS care delivery system and guide policies that will improve the care of older adults with fall injuries. Our aim, therefore, is to estimate the strength of the association between EMS response times and fatal fall injuries among older adults and measure how this association differs by sex.

## Methods

### Study Design and Population

For this retrospective cohort study, we pooled six years of data (2015 to 2020) from the American College of Surgeons’ National Trauma Data Bank (NTDB).^17^ The NTDB is the largest trauma registry in the United States (US), collecting data on trauma-related hospital admissions from over 700 trauma centers across the US.^17^ Each year of privately released data, following an approved request, contains over 3 million de-identified trauma patients managed across trauma-center levels I to V as well as non-designated trauma centers across the US.^17^

### Inclusion and Exclusion Criteria

Across selected years, we identified adults 65 years and older (N=1,796,410) (Figure 1). We limited the data to older adults who presented to an emergency department (ED) with ICD 10 codes W01 -W19 (“Falls”), Y01 (“Assault by pushing from a high place”), and Y30 (“Falling, jumping or pushed from a high place, undetermined intent”) indicating fall-related injuries (N=1,395,024). We excluded patients whose mortality status was not known (n = 116,150; 6.5% of 1,796,410) and patients without recorded EMS response time (n = 374,496; 20.8% of 1,796,410). We also excluded patients whose response time exceeded 60 minutes since such cases are typically associated with unusual circumstances (n=54,606; 3.0% of 1,796,410). For example, an EMS response time of over 6 hours was documented in Alabama on March 3, 2019, but it occurred during a tornado that involved more than 40 locations.^18^ Furthermore, we excluded patients whose trauma center designation was not known (n=144,351; 8.0% of 1,796,410) and whose sex was not recorded (n=70; <0.01% of 1,796,410). The final dataset, therefore, was 705,491 (39.3% of 1,796,410) adults 65 years and older with fall injuries that were transported to levels I to V trauma centers.

**Figure 1:**
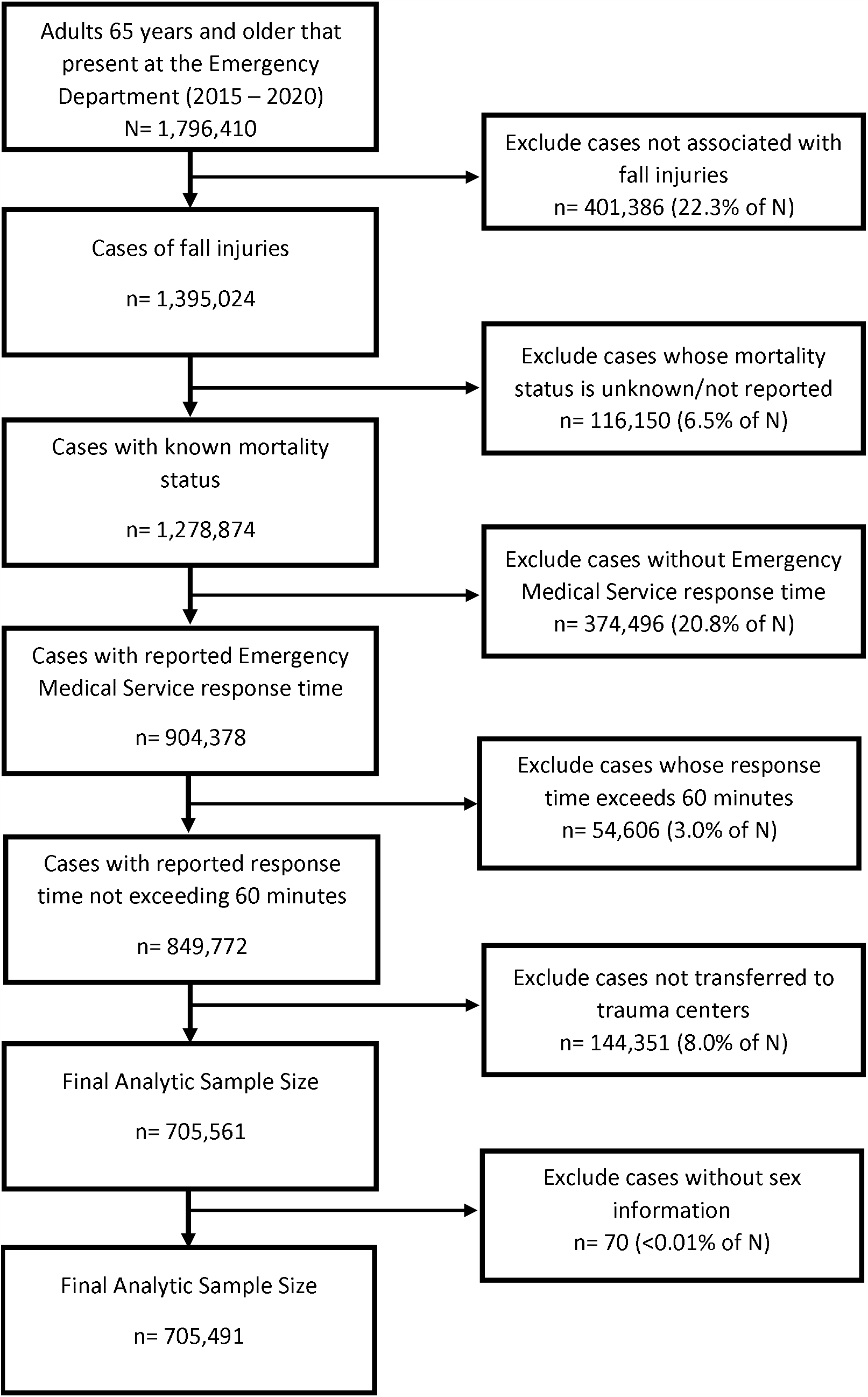
Data selection steps

### Fatal Fall Injury

The outcome variable was fatal fall injury defined as fall-related mortality that occurred during pre-hospital care, ED admission, or during the index hospital stay. Fatal fall injury was measured as a binary variable.

### EMS Response Time

The predictor variable was the EMS response time defined as the EMS travel time – the duration of departure from the base station to the scene of injury. EMS response time was measured as both a continuous measure and as a categorical variable. We created meaningful categories based on the less-than-nine-minute EMS travel time benchmark set by the Fire and EMS Department^19^ and the National Fire Protection Agency.^20^ We defined these categories as follows: Less than nine minutes, nine to 17.59 minutes, 18 to 26.59 minutes, and 27 minutes or higher.

### Trauma Level Designation

We used trauma center level designation as a random effect variable. Trauma center designation is defined as levels I to V. A level I trauma center is a tertiary care facility that provides the highest level of trauma care, serves as a regional trauma facility, and has 24-hour in-in-hospital coverage by general surgeons and surgery specialties.^21, 22^ A level II trauma center has 24-hour in-hospital coverage by a general surgeon but can refer specialized cases such as cardiac and microvascular surgery to level I trauma centers. A level III trauma center has 24-hour in-hospital coverage by emergency physicians, transfers cases to either level I or II trauma centers, and serves as a backup trauma care center for rural and community hospitals. A level IV trauma center provides advanced trauma life support, stabilizes trauma patients, and refers to higher-level trauma centers. A level V trauma center contains basic emergency department facilities and stabilizes patients for transfer to levels I to III trauma centers. The NTDB has no entry for level V trauma center.

### Control Variables

We controlled for age, sex, race/ethnicity, and the presence of coexisting comorbidities. Age was measured as a continuous variable while sex was measured as a binary variable. We defined race/ethnicity as a four categorical level comprising of non-Hispanic Whites, non-Hispanic Blacks, Hispanics, and other races. The comorbidities we measured were diagnoses of hypertension, diabetes, and chronic obstructive pulmonary disease (COPD).

### Data Analysis

We report the frequency distribution and summary statistics (mean, standard deviation (SD), median, and first and third quantile (Q1, Q3)) for categorical and continuous variables as appropriate. We computed the yearly fall case fatality rate by dividing the number of fatal fall counts by the population of older adults with fall injuries for each year. We assessed the statistical significance of differences between fall-related mortality and the patients’ sociodemographic, health, and injury characteristics using the chi-square test, independent sample t-test, and Mann-Whitney U test as appropriate. We report the unadjusted and adjusted odds (plus 95% confidence interval (CI)) of fall-related mortality from a mixed multivariable logistic regression model. For the mixed model, we used the EMS response time, sociodemographic and health characteristics as fixed effect measures, and the trauma level designation as a random effect measure. We also created an interaction model comprising of EMS response time and sex, and we reported the predicted probabilities (plus 95% CI) of these two variables while holding other variables constant at their mean values. Data were analyzed using R (version 4.2.1)/RStudio^23, 24^ and STATA version 17.^25^

## Results

Of 705,941 older adults in the study population, the mean (SD) age was 78.1 (7.1) years (Table 1). The population was predominantly female (60%) and non-Hispanic White (82%). Sixty-eight percent had a diagnosis of hypertension while the proportion of the sample population that had diabetes mellitus and COPD was 26% and 15%, respectively. Most patients were managed in level I (44%) and level II (42%) trauma centers. The median (Q1, Q3) EMS response time was 8 minutes (5.0, 13.0) and 60% of the patients experienced an EMS response time of less than nine minutes.

**Table 1:** Demographic, health, and injury characteristics of older adults with fall injuries (N=705,491).

| Variables | All population (%)<br>N = 705,491 | Fatal Fall<br>n= 31,117 (4.4%) | Non-fatal Fall<br>n=674,374 (95.6%) | p-value |
| --- | --- | --- | --- | --- |
| Age (Mean (SD))<sup>a</sup> | 78.1 (7.1) | 79.2 (6.8) | 78.1 (7.1) | <0.001 |
| Sex |  |  |  |  |
| Female | 423,633 (60.0) | 13,071 (42.0) | 410,562 (60.9) | <0.001 |
| Male | 281,858 (40.0) | 18,046 (58.0) | 263,812 (39.1) |  |
| Race/Ethnicity |  |  |  |  |
| Non-Hispanic Whites | 575,788 (81.6) | 24,736 (79.5) | 551,052 (81.7) | <0.001 |
| Non-Hispanic Blacks | 33,464 (4.7) | 1,438 (4.6) | 32,026 (4.8) |  |
| Hispanics | 34,738 (4.9) | 1,643 (5.3) | 33,095 (4.9) |  |
| Other Races | 26,039 (3.7) | 1,548 (5.0) | 24,491 (3.6) |  |
| Unknown | 35,462 (5.1) | 1,752 (5.6) | 33,710 (5.0) |  |
| Hypertension |  |  |  |  |
| No | 226,557 (32.1) | 10,402 (33.4) | 216,155 (32.0) | <0.001 |
| Yes | 478,934 (67.9) | 20,715 (66.6) | 458,219 (68.0) |  |
| Diabetes Mellitus |  |  |  |  |
| No | 519,782 (73.7) | 21,992 (70.7) | 497,790 (73.8) | <0.001 |
| Yes | 185,709 (26.3) | 9,125 (29.3) | 176,584 (26.2) |  |
| COPD |  |  |  |  |
| No | 602,643 (85.4) | 25,600 (82.3) | 577,043 (85.6) | <0.001 |
| Yes | 102,848 (14.6) | 5,517 (17.7) | 97,331 (14.4) |  |
| Trauma Center Designation |  |  |  |  |
| Level I | 310,322 (44.0) | 16,486 (53.0) | 293,836 (43.6) | <0.001 |
| Level II | 295,169 (41.8) | 12,081 (38.8) | 283,088 (42.0) |  |
| Level III | 97,154 (13.8) | 2,486 (8.0) | 94,668 (14.0) |  |
| Level IV | 2,846 (0.4) | 64 (0.2) | 2,782 (0.4) |  |
| EMS Response time (mins) |  |  |  |  |
| Median (Q1, Q3)<sup>b</sup> | 8.0 (5.0 – 13.0) | 8.0 (5.0 – 15.0) | 8.0 (5.0 – 13.0) | <0.001 |
| EMS Response Categories |  |  |  |  |
| Less than 9minutes | 424,266 (60.1) | 17,573 (56.5) | 406,693 (60.3) | <0.001 |
| 9 to 17.59 minutes | 186,100 (26.4) | 7,874 (25.3) | 178,226 (26.4) |  |
| 18 to 26.59 minutes | 52,630 (7.5) | 3,017 (9.7) | 49,613 (7.4) |  |
| 27 minutes or higher | 42,495 (6.0) | 2,653 (8.5) | 39,842 (5.9) |  |

Test of association was the chi-square test except otherwise specified; a: One-sample t-test performed; b: Mann-Whitney U test performed

The fall case fatality rate was 4.4% and across the study years, the yearly case fatality rate ranged from 4.1% to 4.6% (Figure 2). Among male older adult patients, the fall case fatality rate was 6.4% and the yearly case fatality rate ranged from 6.0% to 6.9%. Among female older patients, the fall case fatality rate was 3.1% and the yearly fall case fatality rate ranged from 2.9% to 3.3%. Fatal fall injury was significantly associated with age, sex, race/ethnicity, hypertension, diabetes mellitus, COPD, and trauma level designation (p<0.001 for all) (Table 1). Fifty-seven percent of older adults who died had an EMS response time of less than 9 minutes, compared to 60% of those who survived (p<0.001).

**Figure 2:**
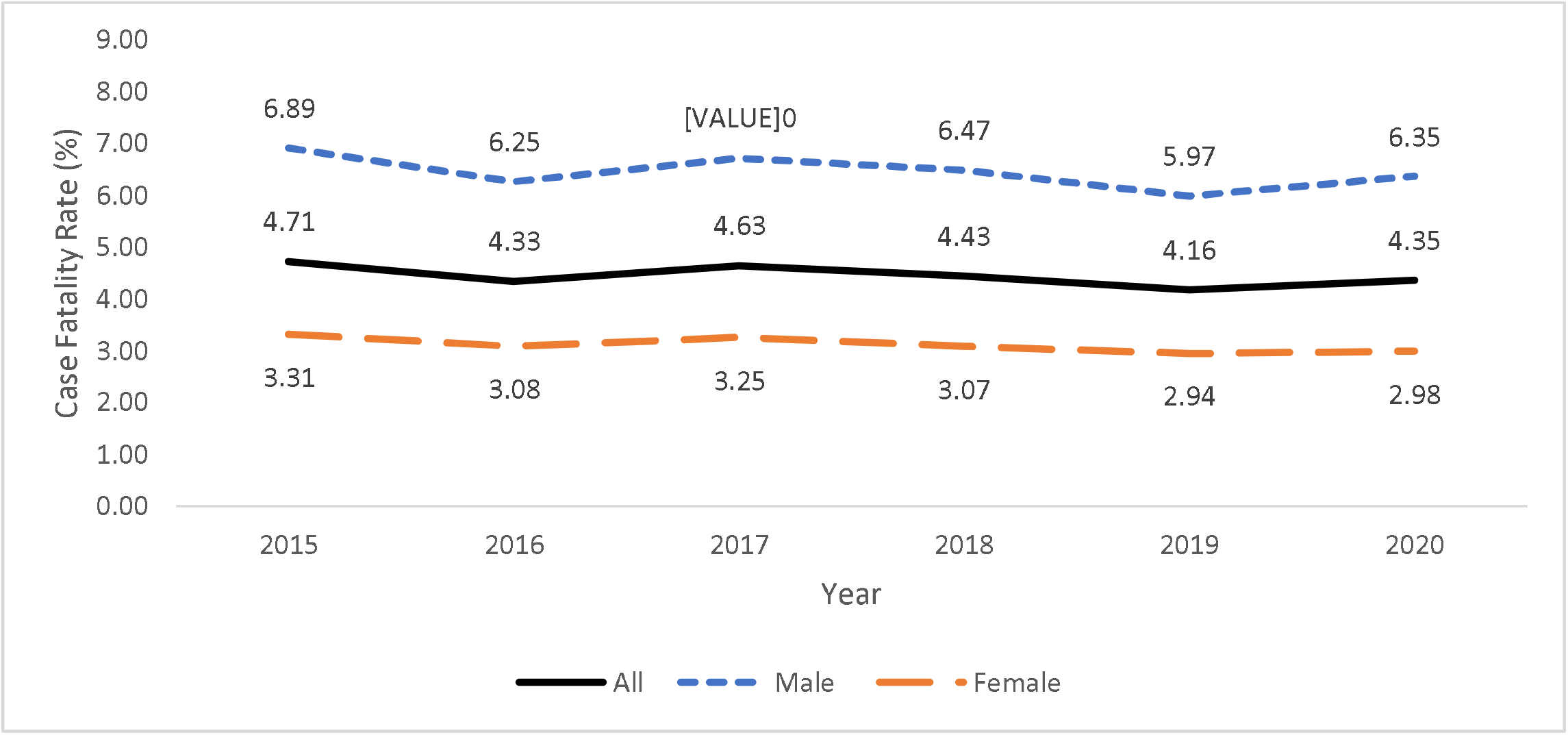
Yearly case fatality rates of fall injuries stratified by male and female older adults.

In the unadjusted model, age, sex, being Hispanic or of other races, and having diagnoses of hypertension, diabetes, or COPD were associated with fatal fall injuries (Table 2). In the adjusted model (Model 1), males had 2.2 times the odds of fatal fall injuries compared to females (OR: 2.18; 95% CI: 2.05 - 2.32). Also, a minute increase in EMS response time was associated with a 1% increased odds of fatal fall injury (OR: 1.01; 95% CI: 1.01 - 1.01) (Model 1). Compared to patients who experienced EMS response time of less than nine minutes, those who experienced EMS response of 18 to 26.59 minutes, and 27 minutes or higher had 33% (Adjusted OR (AOR): 1.33; 95%: 1.28 - 1.39) and 41% (AOR: 1.41; 95%: 1.35 - 1.47) increased odds of fatal fall injuries (Model 2). In the interaction model, EMS response times of 9 to 17.59 minutes, 18 to 26.59 minutes, and 27 minutes or higher were associated with 1.1 (AOR: 1.06; 95% CI: 1.01 - 1.10), 1.4 (AOR: 1.37; 95% CI: 1.29 - 1.46), and 1.5 (AOR:1.52; 95% CI: 1.42 - 1.62) times the odds of fatal fall injuries, respectively.

**Table 2:**
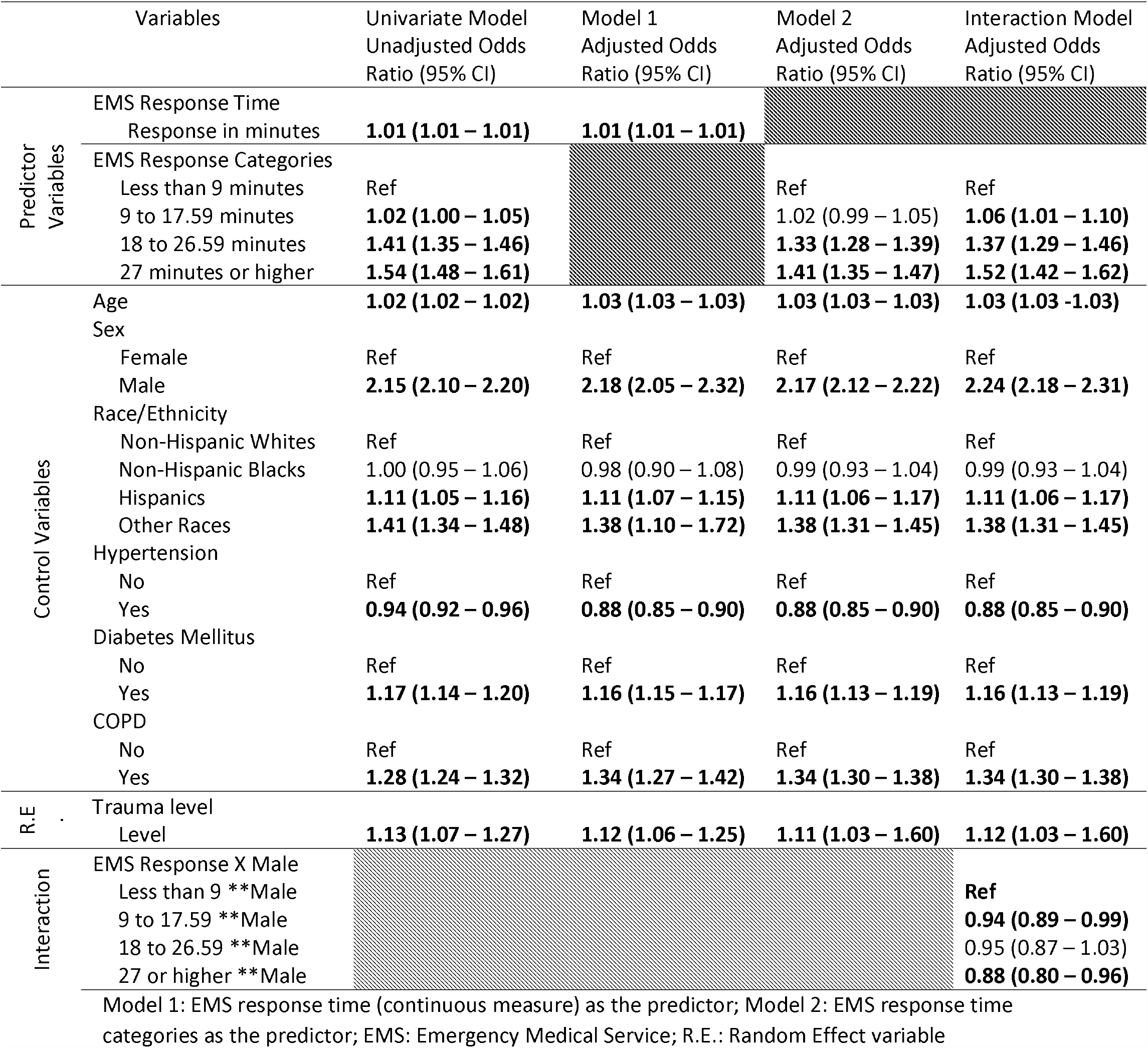
Unadjusted and adjusted odds of fatal fall injuries among older adults (N=705,491)

The predicted probability of fatal fall injury was 3.3% (95% CI: 2.22 – 4.35) while the predicted probability of fatal fall injury among male and female older adults was 5.1% (95% CI: 3.51 – 6.76) and 2.4% (95% CI: 1.64 – 3.22), respectively (Figure 3). Among male older adults, the predicted probability of fatal fall injury was lowest at 5.0% (95% CI: 3.40 – 6.54) when the EMS response time was less than nine minutes and the predicted probability increased to a high of 6.5% (95% CI: 4.46 – 8.54) when EMS response time was 27 minutes or higher.Among female older adults, the predicted probability of fatal fall injury was lowest at 2.3% (95% CI: 1.54 – 3.03) when the EMS response time was less than nine minutes and the predicted probability increased toa high of 3.4% (95% CI: 2.30 – 4.55) when EMS response time was 27 minutes or higher.

**Figure 3:**
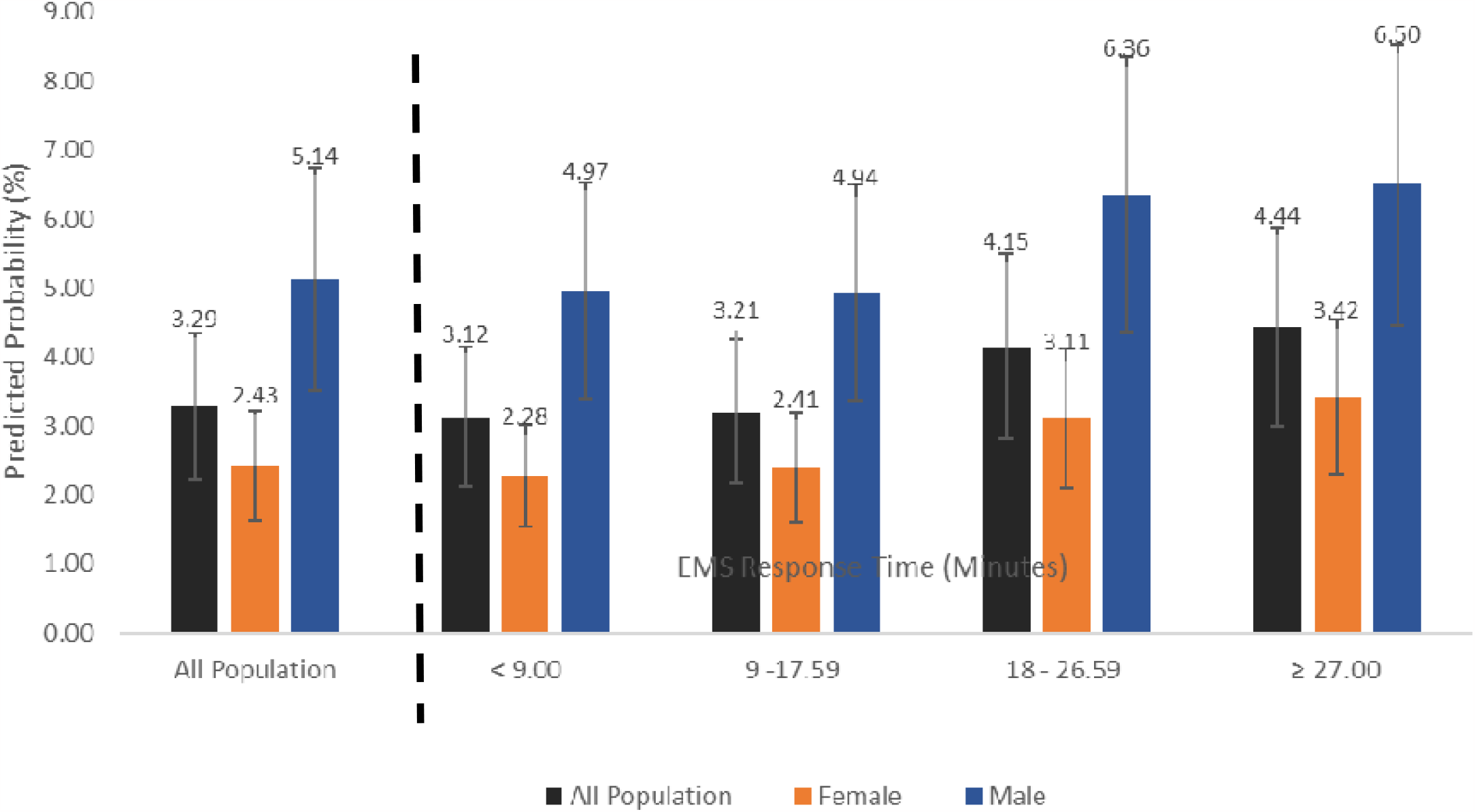
Predicted probabilities of fatal fall injuries among older adults across EMS response categories

## Discussion

To our knowledge, this nationally representative cross-sectional study is the first to report the dose-response relationship between EMS response time and fatal fall injuries among older adults. Additionally, this study adds to the existing fatal fall injury epidemiology by reporting the fall-related case fatality rates among older US adults who were transported by the EMS to trauma centers. While earlier studies have reported population-based age-standardized geriatric fatal fall rates of 62 per 100,000,^1, 4^ we have provided hospital-based case fatality rates that show that four of every 100 older adults transported by the EMS to trauma centers die from fall injuries.

Earlier studies have reported the association between advanced age, frailty, co-morbid conditions, and fall-related mortality.^5, 26-28^ All of these indicators are essentially non-modifiable. Our study identifies an important modifiable factor by establishing EMS response time as a predictor of fall-related mortality in older persons. Rather than translating this result into a call for increased driving speed by ambulance drivers, we believe a greater acceptance and increased use of technology in EMS delivery can shave precious minutes off response times and result in fewer deaths. Medical drones, for example, can shorten travel time and complement EMS workers in delivering care, especially in areas where achieving the EMS response benchmark is logistically impossible.^29^ Currently, medical drones can transport medications and blood, engage in search and rescue missions, transport patients, and provide telemedicine services.^30^ Increased acceptance, utilization, and integration of medical drones into EMS will encourage further adaptation of the device for additional acute and critical care-related tasks.

Among older adults with fall injuries, the median EMS response time was eight minutes and approximately 60% of these patients experienced a less-than-nine-minute EMS response time. The NFPA Standard 1710 had set a benchmark of less than eight minutes 90% of the time for cases with low to medium hazard risk and less than 10 minutes 90% of the time for cases with high hazard risk.^19, 31^ While EMS has achieved these standards with a median response time of 8 minutes, this benchmark was met 60% of the time. Achieving the eight-minute or ten-minute benchmark 90% of the time may require a data-driven approach to spatially optimize routes and coverage areas of current EMS stations, and identify the need and best location for additional EMS stations.^32-34^ Concerted efforts by government, community stakeholders, and EMS administrators to fund the EMS infrastructure, train workers, and educate the population in communities with the greatest risk of fatal fall injuries may be needed for success.

This study is subject to a number of important limitations. As a cross-sectional study, causal inferences are not possible. While the NTDB is the largest trauma data pool across the US, it is not a census of all US trauma cases and is not necessarily broadly representative of the entire US trauma population. We did not control for rurality/urbanicity since this measure was not provided in the NTDB data released for this study. However, earlier studies had reported longer median EMS response times in rural areas, with 33% and 46% of crash victims experiencing less than nine minutes of EMS response time.^9, 11^ It is, therefore, likely that a similar pattern would be present among older adults with fall injuries in rural areas. Furthermore, we controlled for trauma center designation, a variable that is strongly associated with rurality/urbanicity.^35^ Level I and II trauma centers are often inaccessible in rural and underserved areas where levels III to IV predominate.^36^ Also, the absence of county identifiers limits the generalization of the results for policy recommendations and community interventions. We used data that captured the pre-COVID (2015 to 2018) and intra-COVID (2019 - 2020) periods. Earlier studies had reported reduced hospital utilization, less EMS dispatch, and more out-of-hospital deaths for non-COVID cases.^37-39^ Our computed fall case fatality rates may be conservative estimates and the fatality rates may be much higher in the general population during the COVID period. Also, with data entry, errors may result in a misclassification bias. Our results would be conservative if such bias is present. Misclassification of outcomes, however, is unlikely since death is a terminal diagnosis.

## Conclusion

This is, to our knowledge, the first study of the relationship between EMS response time and fatal fall injury mortality among older adults in the US. Delay in EMS response time is associated with fatal fall injuries. Improving the EMS infrastructure, especially in communities that are logistically impossible to meet the national benchmark, may reduce the odds of fatal fall injuries among older adults.

## Data Availability

All data produced in the present study are available upon reasonable request to the authors

## Acknowledgment

The authors appreciate the Committee on Trauma, American College of Surgeons for providing the National Trauma Data Bank for use.

Committee on Trauma, American College of Surgeons. NTDB Admission Year 2015 - 2020 Chicago, IL.

The content reproduced from the applications remains the full and exclusive copyrighted property of the American College of Surgeons. The American College of Surgeons is not responsible for any ancillary or derivative works based on the original data, text, tables, or figures.

